# Body mass index and the duration of the pubertal growth spurt in Vietnamese urban children

**DOI:** 10.64898/2026.09.09.26362696

**Authors:** Nhan T. Ho

**Author notes:** Corresponding authors: Nhan T. Ho, Research Management Department, Vinmec International Hospital, Hanoi, Vietnam and College of Health Sciences, VinUniversity, Hanoi, Vietnam.

## Abstract

**Background:** Higher childhood body mass index (BMI) is linked to earlier and less intense pubertal growth spurts, but whether it also changes the spurt duration is rarely studied, especially in Vietnam and Southeast Asia.

**Methods:** We used previously fitted Superimposition by Translation and Rotation (SITAR) growth curves for 8,018 boys and 5,491 girls in a multicity Vietnamese cohort. For each child we derived two duration measures from their own predicted velocity curve, a full width at half maximum (FWHM) measure and a threshold free growth quartile duration, and tested associations with prepubertal BMI z score before and after adjusting for pubertal timing and peak height velocity.

**Results:** FWHM was identifiable in 99.9% of boys but only 18.0% of girls, and identifiable girls had systematically lower BMI, indicating informative missingness. Growth quartile duration was available for nearly all boys and 94.9% of girls. Higher BMI predicted shorter unadjusted duration in both sexes (p < 0.001 to p = 0.037). After adjustment, in boys, the association weakened but stayed significant. In girls, growth quartile duration reversed direction and remained significant (0.0052 years per unit BMI z score, p = 0.004), with a significant BMI by sex interaction (p = 0.007). Functional principal component analysis confirmed these findings independent of any single model parameter.

**Conclusion:** Pubertal growth spurt duration responds to BMI differently by sex once timing and intensity are accounted for. Full width based duration measures should be checked against each child’s own available age range before use, particularly in girls.

## Introduction

Puberty is when children add most of the height they will carry into adulthood, and how that growth unfolds has drawn growing attention as childhood overweight and obesity have risen worldwide. Height and body mass index trajectories through childhood and adolescence vary widely across countries, reflecting differences in nutrition and living conditions with consequences that persist into adult life ^1^.

Body mass index before puberty is one of the more consistent predictors of how puberty unfolds. Cohort studies from China and the United States show that children with higher prepubertal body mass index, or faster weight and length gain in early life, tend to reach puberty and their age at peak height velocity earlier than leaner children ^2,3^. This earlier timing does not translate into a bigger pubertal growth spurt. In Chinese, Swedish, and mixed Belarusian and American cohorts, higher body mass index is associated with more growth before puberty begins but a smaller and slower pubertal height gain once it starts, so heavier and leaner children can still reach similar adult heights through very different paths ^2,4,5^. Among children with severe obesity, this blunting of the pubertal growth spurt is even more pronounced ^6^. The pattern is not universal. In a Taiwanese cohort, puberty itself outweighed body mass index as the main driver of growth velocity in girls entering adolescence, pointing to sex specific modulation around the spurt ^7^.

The method most often used to quantify these patterns is SITAR, Superimposition by Translation and Rotation, which fits each child’s own growth curve as a shifted and scaled version of a shared population curve ^8^. SITAR separates a timing parameter that shifts the curve earlier or later in age from an intensity related parameter that stretches or compresses it. This framework has been applied to pubertal height growth in a trans ancestry genetic study, a multinational cohort of adolescents with perinatal HIV, and the pace of change in body composition rather than height alone ^9–12^.

What none of these applications separates cleanly from intensity is duration. Because that parameter rescales the age axis as a whole, a more intense spurt is, by the mathematics of the model, also a more compressed one. Whether body mass index changes how long the pubertal growth spurt lasts, apart from how fast or how early it happens, is a related but distinct question that has drawn far less attention. One Chinese cohort came closest, defining duration from growth takeoff to peak height velocity as its own parameter and linking a shorter pubertal height spurt to higher risk of overweight and obesity in late adolescence ^13^. A Danish cohort described pubertal tempo as a further dimension beyond timing, finding that faster tempo remained associated with body mass index in young adulthood even after accounting for childhood body mass index, suggesting tempo is not simply a byproduct of timing or of childhood weight ^14^.

Beyond these two studies, duration or tempo of the pubertal growth spurt is rarely estimated as an outcome in its own right, and we are not aware of a measure built directly from the shape of a child’s own velocity curve rather than a single model parameter tied mathematically to intensity.

This gap is wider still for Vietnam and Southeast Asia. Vietnamese girls were among the countries with more favorable long-term height and body mass index patterns in a recent global comparison, though Vietnam has rarely been the focus of a dedicated cohort study of pubertal growth ^1^. A multinational cohort of adolescents with perinatal HIV found that those in the Asia Pacific region had shorter stature and later, less intense growth spurts than peers in Europe or North America ^10^. Beyond this indirect signal and the Taiwanese evidence noted above ^7^, no Vietnamese or Southeast Asian cohort, to our knowledge, has directly tested whether body mass index is associated with pubertal growth spurt duration.

We addressed this gap using longitudinal height data from a multicity Vietnamese schoolchildren cohort with SITAR growth models for boys and girls fitted in our own earlier work ^15,16^.

Building on that work, which described pubertal timing and the distribution of peak height velocity by body mass index in this cohort, we asked a further question, whether prepubertal body mass index associates with the pubertal growth spurt duration, using a duration measure derived directly from each child’s own velocity curve rather than a single SITAR parameter.

## Methods

### Study population and prior analyses

This study performed retrospective analysis of de-identified annual school health check data of children attending a private school system in 3 major cities in Vietnam (Hanoi, Ho Chi Minh City, and Haiphong) between 2018 and 2025. This study was approved by Vinmec Ethical Committee (approval number 0231/2024/CN/HDDD VMEC). A waiver of individual informed consent was granted as the study used de-identified routinely collected retrospective health data from school health examinations.

The study, all methods and report were carried out in accordance with the Declaration of Helsinki and STROBE (STrengthening the Reporting of OBservational studies in Epidemiology) guideline. STROBE checklist with our study corresponding status was included in Supplementary Note.

Cohort assembly, anthropometric quality control, and World Health Organization reference standards for body mass index z-scores ^17^ have been described in earlier papers from this series ^15,16^. The present paper builds directly on the fitted SITAR (Superimposition by Translation and Rotation) growth models reported previously, one for boys spanning ages 6 to 18 years and one for girls spanning ages 6 to 16 years, selected through formal model comparison ^8^. These models were not refit here. Instead, we reused the saved model objects, individual random effects, and population parameters for age and peak height velocity at adolescent peak height velocity (APHV and PHV).

Because the fitted female model showed a mild boundary artifact in its predicted velocity curve beyond a data density threshold, we restricted all female velocity evaluations to ages at or below this threshold, following the same convention used in earlier papers in this series.

### Exposure

The exposure of interest was prepubertal body mass index, expressed as a z-score relative to the WHO growth reference and measured before the onset of the pubertal growth spurt for each child. Children were also classified into four categories, thinness, normal weight, overweight, and obesity, using standard WHO cutoffs.

### Deriving individual velocity curves and duration measures

For each child, we predicted height at ages spaced one month apart on either side of that child’s own APHV, then computed height velocity by central finite differences. This produced a dense, individual velocity curve aligned in developmental time rather than chronological age, extending up to five years before and after APHV where the fitted model’s age range allowed it.

From each child’s velocity curve we derived two duration measures. The first was full width at half maximum, the age span during which velocity remained at or above 25, 50, or 75% of that child’s own peak velocity, located by linear interpolation between grid points. The second was a threshold free growth quartile duration, defined as the age interval between the 25th and 75th percentile of cumulative height gained across the observed window, computed by trapezoidal integration of the velocity curve. Because girls’ fitted age range spans roughly 8.5 years compared with about 12 years for boys, each child’s growth quartile duration was flagged as based on a truncated window whenever her or his achieved data fell short of what was actually achievable given that child’s own APHV and sex specific age bounds, rather than short of a single fixed target shared by both sexes. Truncated values were excluded from primary analysis.

### Statistical analysis

Associations between prepubertal body mass index and each duration measure were estimated separately by sex using linear regression with heteroskedasticity consistent standard errors of the HC3 type ^18,19^, adjusting for study city. We complemented this with quantile regression across the duration distribution, fit using the Frisch Newton interior point algorithm for computational efficiency at this sample size ^20,21^, with confidence intervals from 300 city stratified bootstrap resamples ^22^.

To test whether body mass index predicts duration independent of pubertal timing and intensity, we fit an adjusted model including each child’s SITAR timing parameter and individual peak velocity alongside body mass index. We also tested a body mass index by sex interaction term with bootstrap based inference. Sensitivity analyses repeated the primary model across alternative full width thresholds, alternative window widths, and after excluding each study city in turn.

As a threshold free robustness check, we applied functional principal component analysis to each child’s aligned velocity curve using the PACE method ^23^, implemented in the fdapace package ^24^, restricted to a narrower common grid to accommodate girls’ shorter available range. We identified the retained component most strongly correlated with growth quartile duration and regressed its score on body mass index using the same robust standard error approach described above.

All analyses were conducted in R ^25^ using the sitar ^8^, quantreg ^26^, fdapace ^24^, and sandwich ^27^ packages.

## Results

The final analytic sample included 8,018 boys and 5,491 girls with a valid prepubertal body mass index and a fitted SITAR growth curve (**Table 1**). Boys showed a clear gradient across body mass index categories, with an earlier age at peak height velocity and a higher individual peak height velocity as body mass index rose, from 12.43 years and 9.08 centimeters per year in the thinness category to 12.08 years and 9.53 centimeters per year in the obesity category (both p < 0.001). Girls showed the same pattern for peak height velocity, rising from 6.80 to 7.27 centimeters per year across categories (p < 0.001), but age at peak height velocity did not differ meaningfully by body mass index category in girls (p = 0.420).

**Table 1.**
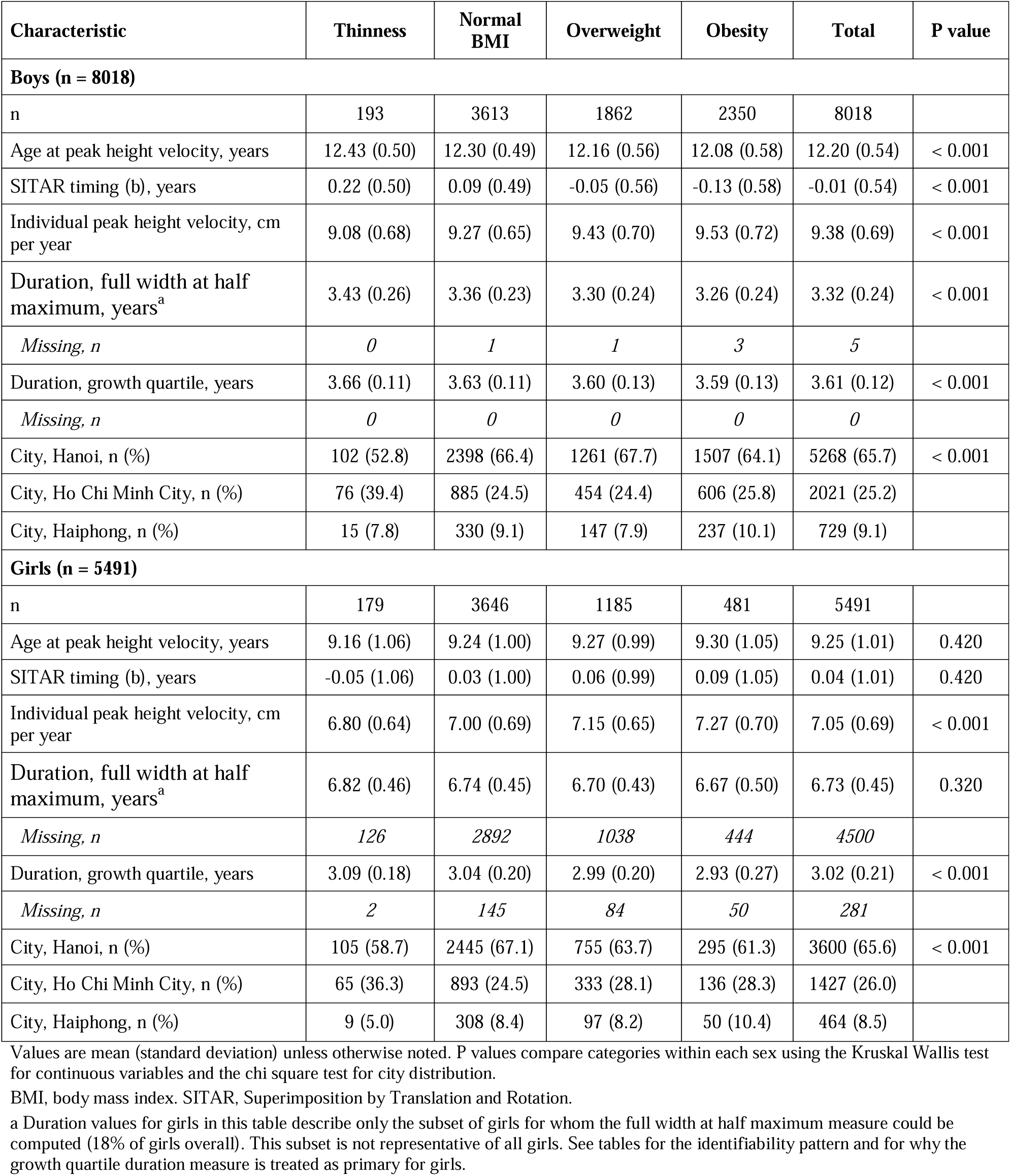
Cohort characteristics by sex and prepubertal body mass index category.

The full width at half maximum duration measure could be computed for nearly all boys, 8,013 of 8,018 (99.9%), but for only 991 of 5,491 girls (18.0%) (**Table S1**). The dominant reason was that the curve did not drop back to half of peak velocity before the peak within the age range supported by the female growth model, affecting 3,513 girls (**Table S2**). Girls with an identifiable duration differed sharply from those without one. The 991 identifiable girls had a mean prepubertal body mass index z score of 0.03 and an obesity prevalence of 3.7%, while the 3,513 girls censored on the pre-peak side had a mean z score of 0.78 and an obesity prevalence of 11.5% (**Table S2**). This pattern, visible directly in the compressed, ceiling bound distribution of female full width at half maximum values across every body mass index category in **Figure 2**, means the full width at half maximum measure is not missing at random in girls and should be interpreted with caution. The threshold free growth quartile duration measure was far more complete, available for 8,018 of 8,018 boys and 5,210 of 5,491 girls (94.9%) (**Table 1**).

**Figure 1.**
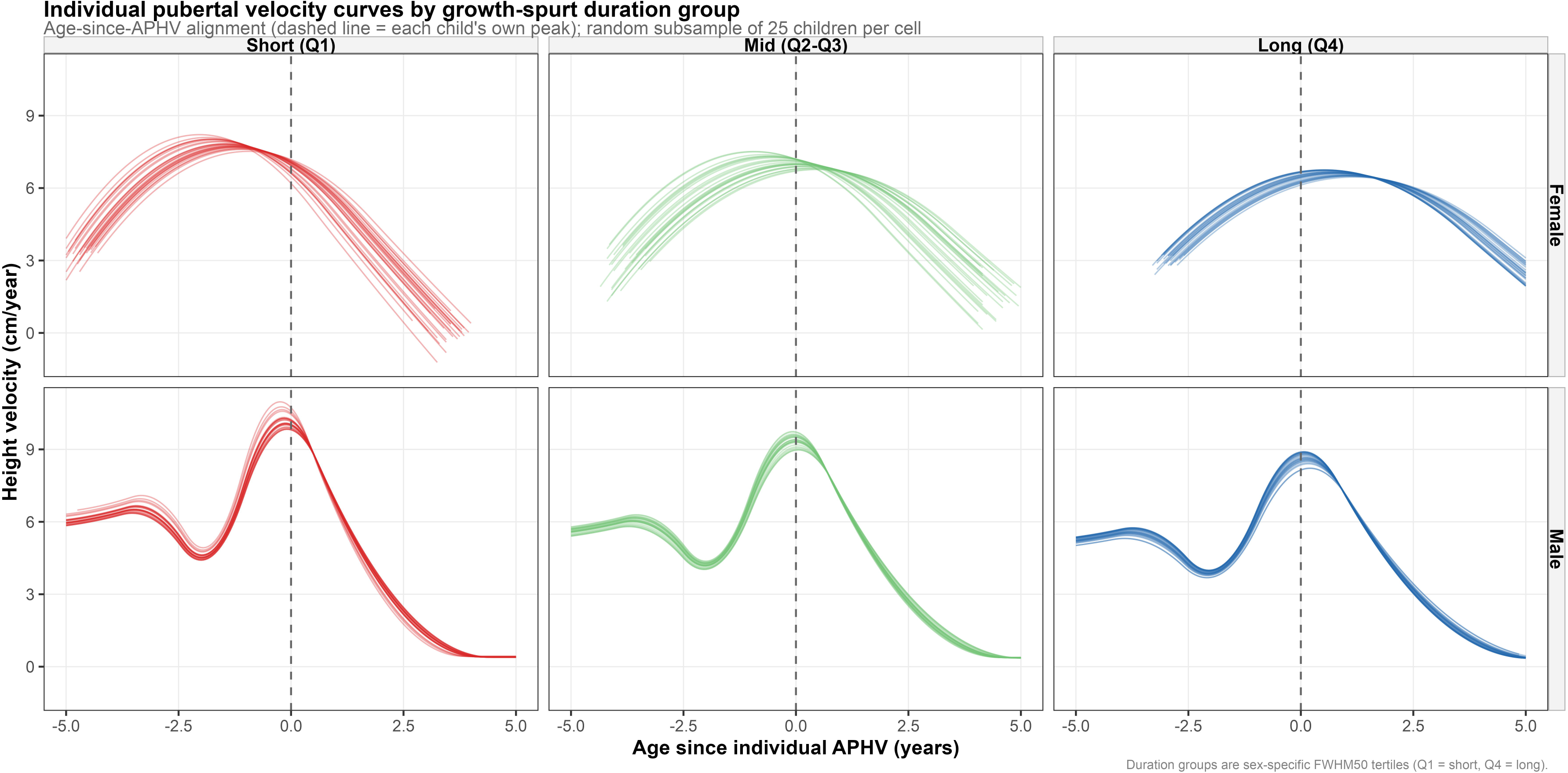
Individual pubertal velocity curves by growth spurt duration group. Each panel shows a random sample of 25 children’s own SITAR predicted height velocity curves, aligned by age relative to that child’s own age at peak height velocity (dashed vertical line at zero). Columns group children into sex specific tertiles of the full width at half maximum duration measure, short (bottom third), mid (middle third), and long (top third). Rows separate girls (top) and boys (bottom). Boys’ curves show the full range from 5 years before to 5 years after peak, including the pre-pubertal deceleration before the pubertal rise. Girls’ curves are shown over a narrower range because of the shorter age span supported by the female growth model.

**Figure 2.**
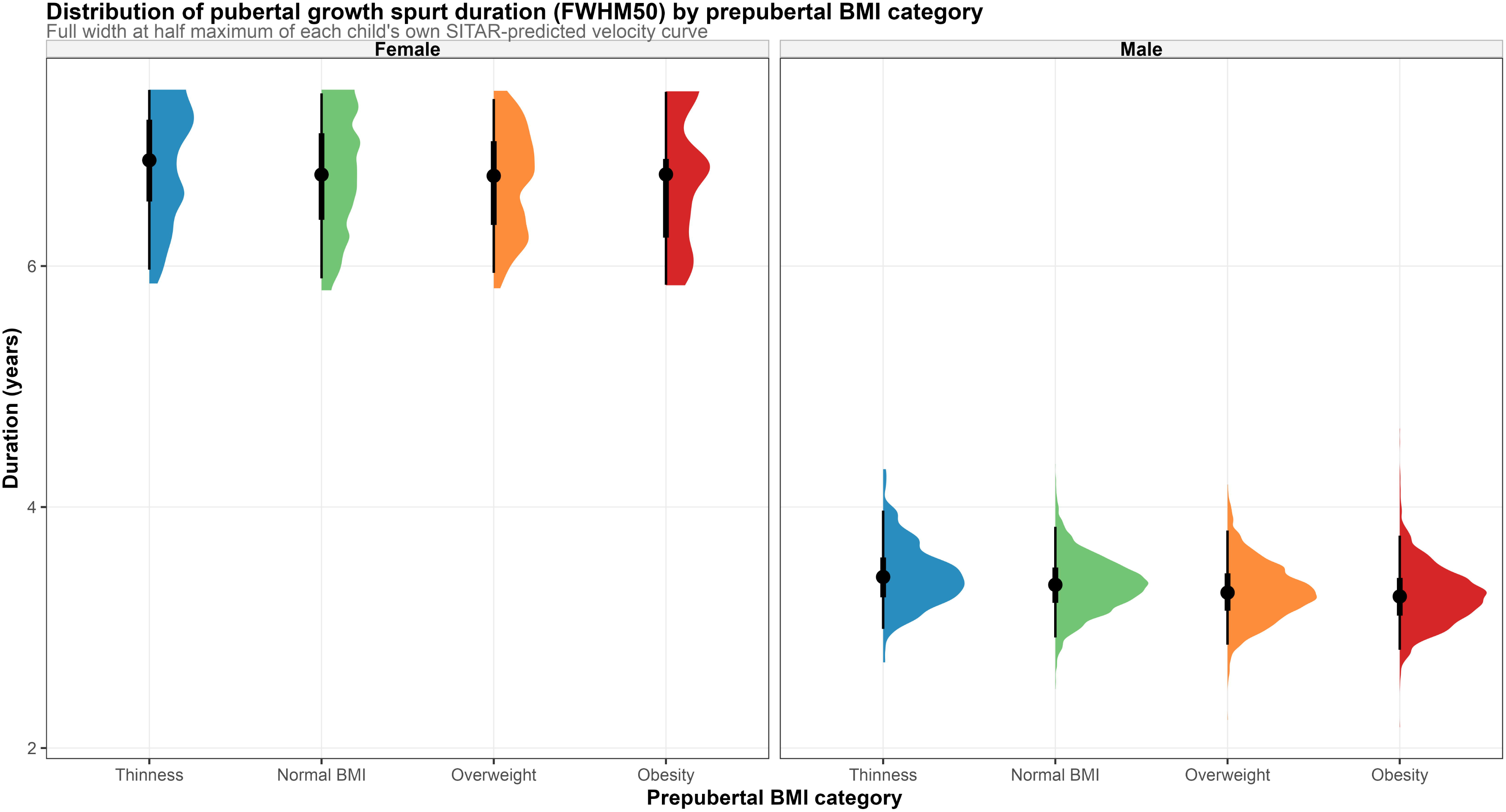
Distribution of pubertal growth spurt duration (full width at half maximum) by prepubertal body mass index category. Half eye plots show the distribution of the full width at half maximum duration measure within each prepubertal body mass index category, separately for girls (left) and boys (right). The dot marks the median and the vertical bar spans the interquartile range. Note that the female distributions cluster tightly near the top of the plotted range in every body mass index category. This reflects the identifiability limitation described in the tables, only girls whose growth spurt happens to fall within their shorter available measurement window can be measured at all, which compresses the observed female values toward the upper limit of what is measurable and away from the fuller range seen in boys. This measure should not be compared at face value between girls and boys.

Duration was correlated with pubertal timing and intensity but was not redundant with either. In boys, full width at half maximum correlated with timing at r = 0.85 and with intensity at r = - 0.99, while timing and intensity correlated with each other at r = -0.85 (**Table 2**). In girls, the corresponding correlations were r = -0.95, r = -1.00, and r = 0.95. The two duration measures agreed closely with each other, at r = 0.95 in boys and r = 0.92 in girls.

**Table 2.**
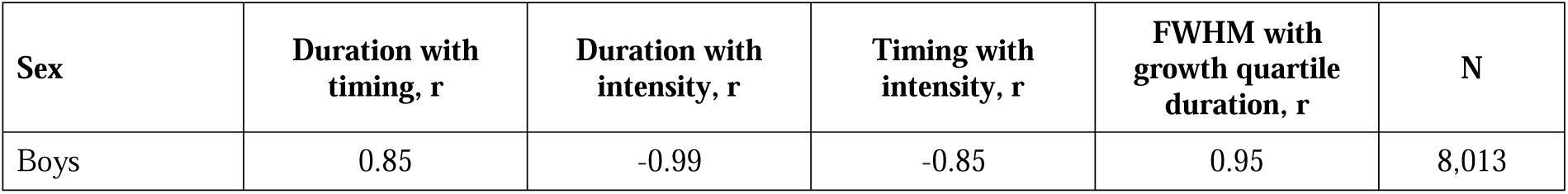
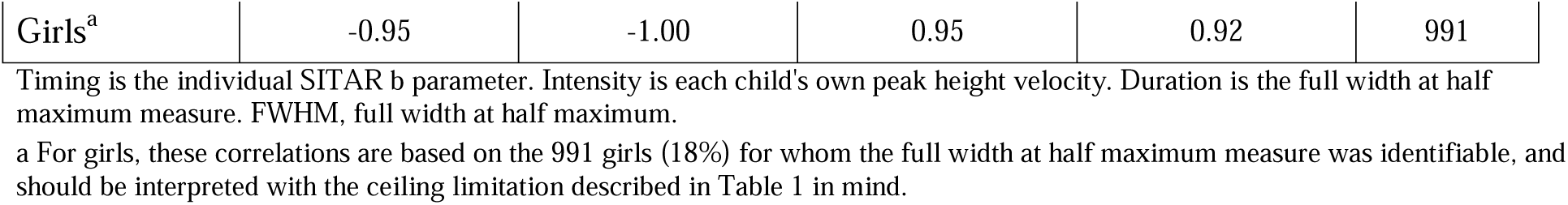
Correlation of pubertal growth spurt duration with pubertal timing and intensity.

Higher prepubertal body mass index was associated with a shorter pubertal growth spurt in the unadjusted models for both duration measures and both sexes (**Table 3**, **Figure 3**). For growth quartile duration, each one unit increase in body mass index z score was associated with 0.0111 fewer years of duration in boys and 0.0278 fewer years in girls (both p < 0.001). After adjusting for pubertal timing and individual peak height velocity, the boys association remained statistically significant but much smaller (-0.0005 years, p = 0.034). In girls, the adjusted association reversed direction and remained significant (0.0052 years, p = 0.004), indicating that once timing and intensity are held constant, higher body mass index is associated with a longer, not shorter, growth quartile duration. The body mass index by sex interaction for growth quartile duration was significant (p = 0.007), confirming that the association genuinely differs between boys and girls rather than reflecting sampling noise. The full width at half maximum results followed a similar unadjusted pattern in both sexes, but neither sex reached significance after adjustment, boys p = 0.038 with a coefficient near zero, girls p = 0.330, and the body mass index by sex interaction for this measure was not significant (p = 0.824).

**Figure 3.**
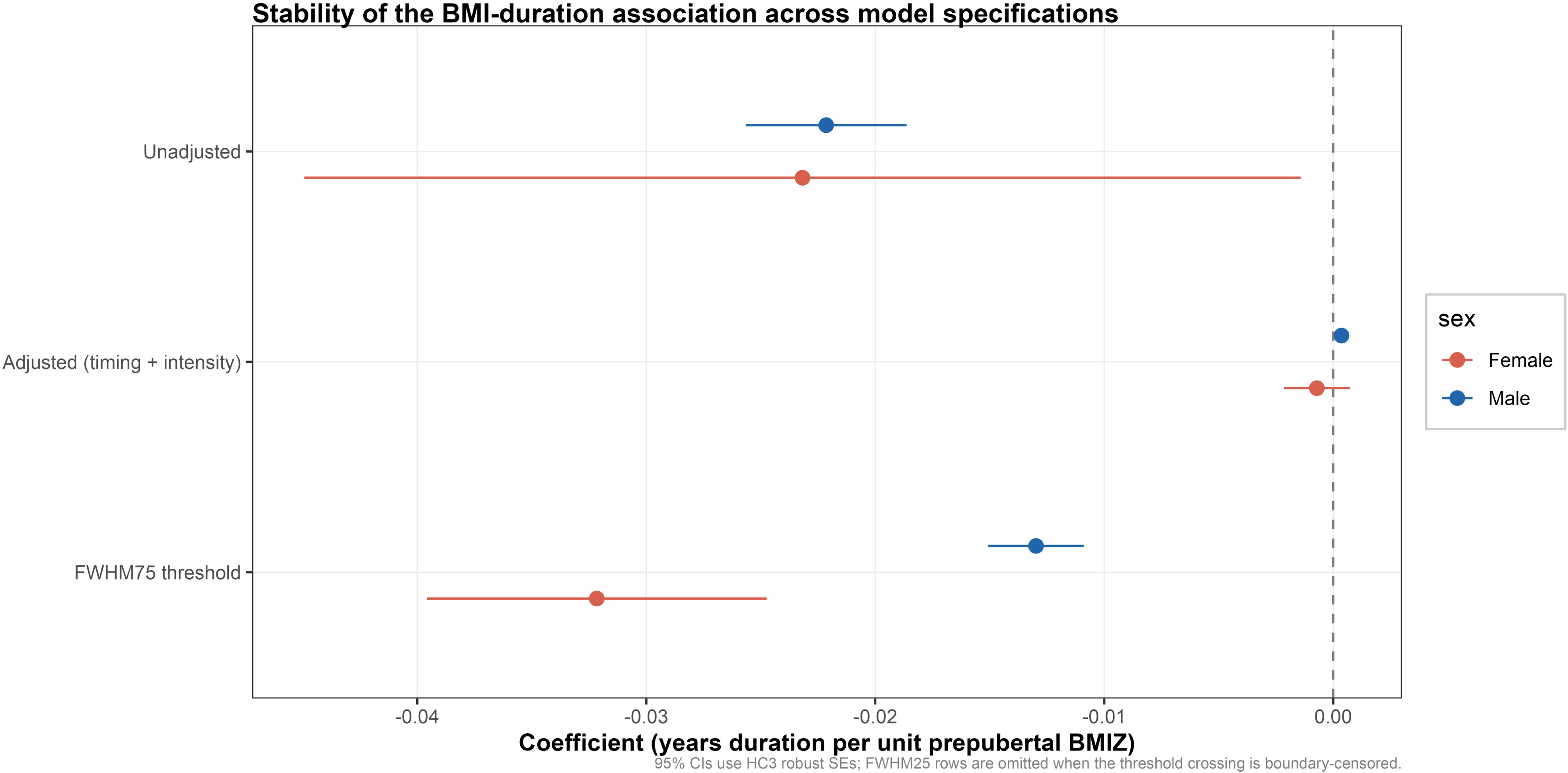
Stability of the body mass index and duration association across model specifications. Points show the coefficient for prepubertal body mass index z score predicting the full width at half maximum duration measure, with horizontal lines showing 95% confidence intervals from heteroskedasticity consistent standard errors. Rows compare the unadjusted model, the model additionally adjusted for pubertal timing and intensity, and the unadjusted model repeated at the looser 75% of peak velocity threshold. The stricter 25% threshold could not be estimated for either sex and is omitted. The dashed vertical line marks no association.

**Table 3.**
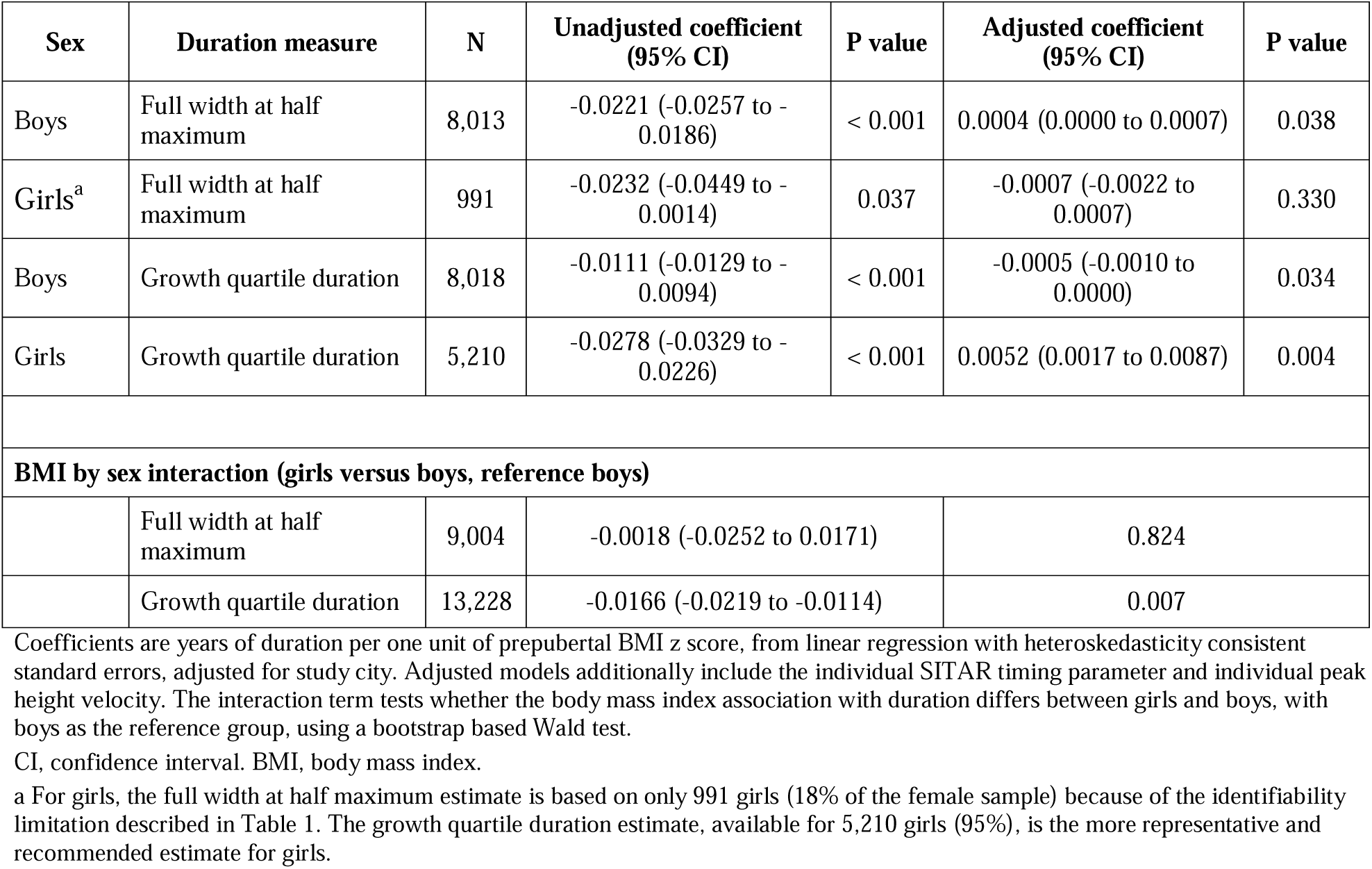
Association between prepubertal body mass index and pubertal growth spurt duration.

The functional principal component analysis provided a threshold free confirmation of these findings. A single retained component explained more than 99% of the variance in the shape of aligned velocity curves in both sexes and correlated strongly with growth quartile duration, r = -0.96 in boys and r = -0.78 in girls, and with full width at half maximum, r = -0.995 and r = -0.992 (**Table 4**, **Figure 4**). This component score was significantly associated with body mass index in both boys and girls (both p < 0.001), reinforcing that the body mass index and duration association is not an artifact of either duration definition.

**Figure 4.**
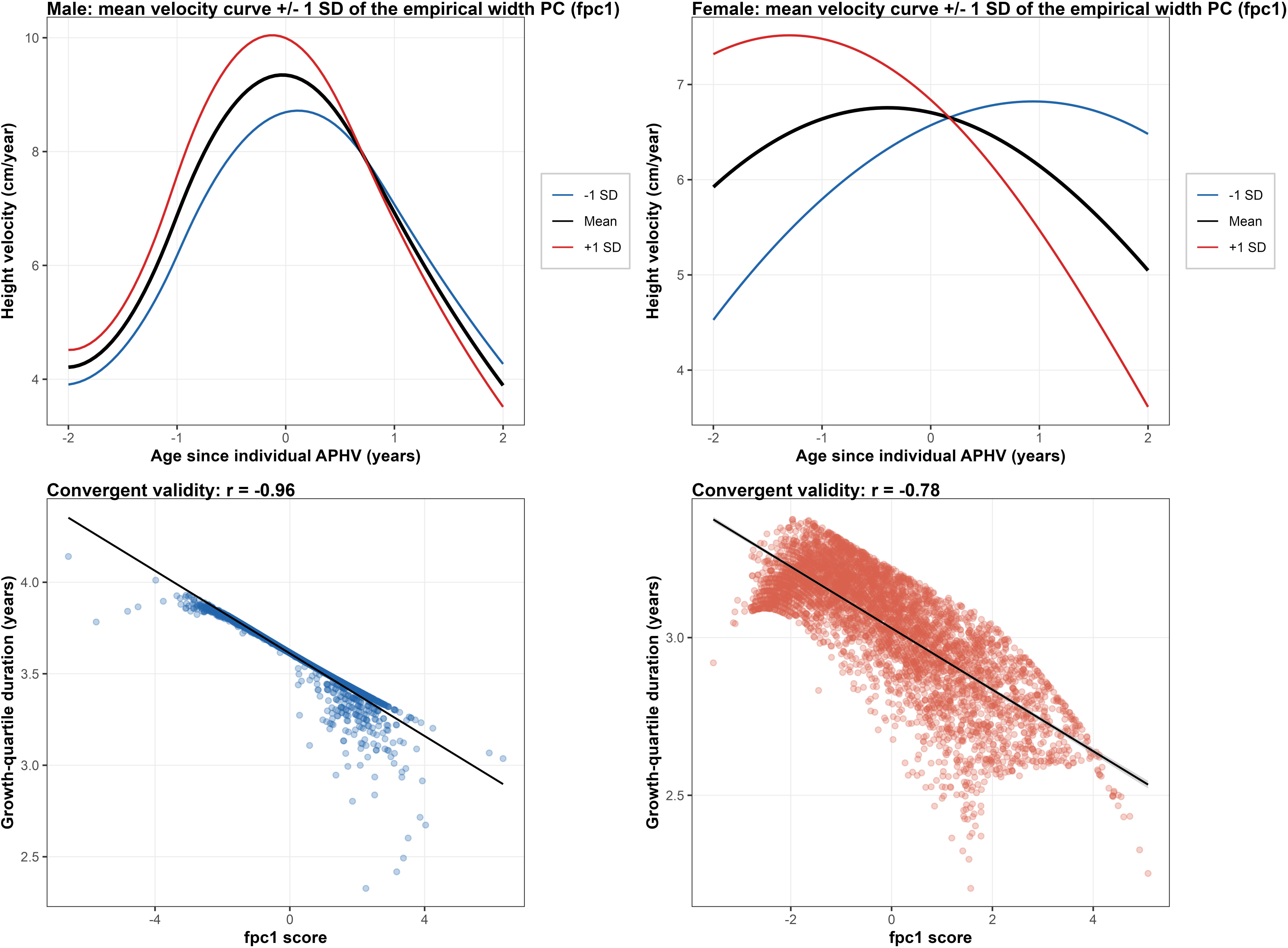
Functional principal component analysis of pubertal velocity curves. Top row, the mean aligned velocity curve (black) for boys (left) and girls (right) within a window of plus or minus 2 years around each child’s own age at peak height velocity, together with the curve shape one standard deviation above (red) and below (blue) the mean on the single retained functional principal component. Bottom row, each child’s score on this component plotted against that child’s growth quartile duration, with a fitted regression line. This component captures the width or spread of the velocity curve directly from the curve shape, without assuming any particular threshold, and its strong correlation with growth quartile duration in both sexes supports the validity of that duration measure.

**Table 4.**
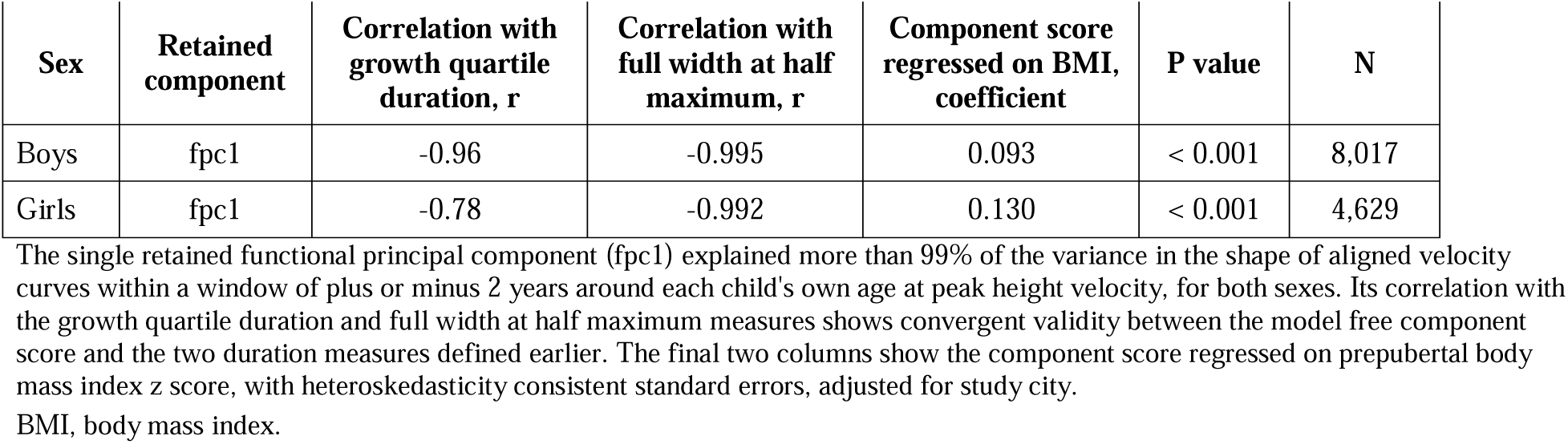
Functional principal component analysis of pubertal velocity curves, robustness check for growth spurt duration.

Quantile regression showed that the growth quartile duration association strengthened toward the shorter end of the distribution in both sexes (Wald p < 0.001 for heterogeneity across quantiles in both boys and girls) (**Table S3, Figure 5**). The full width at half maximum association was stable across quantiles in boys (Wald p = 0.313) but showed no consistent pattern in girls (Wald p = 0.066), consistent with the smaller and less representative female sample for this measure.

**Figure 5.**
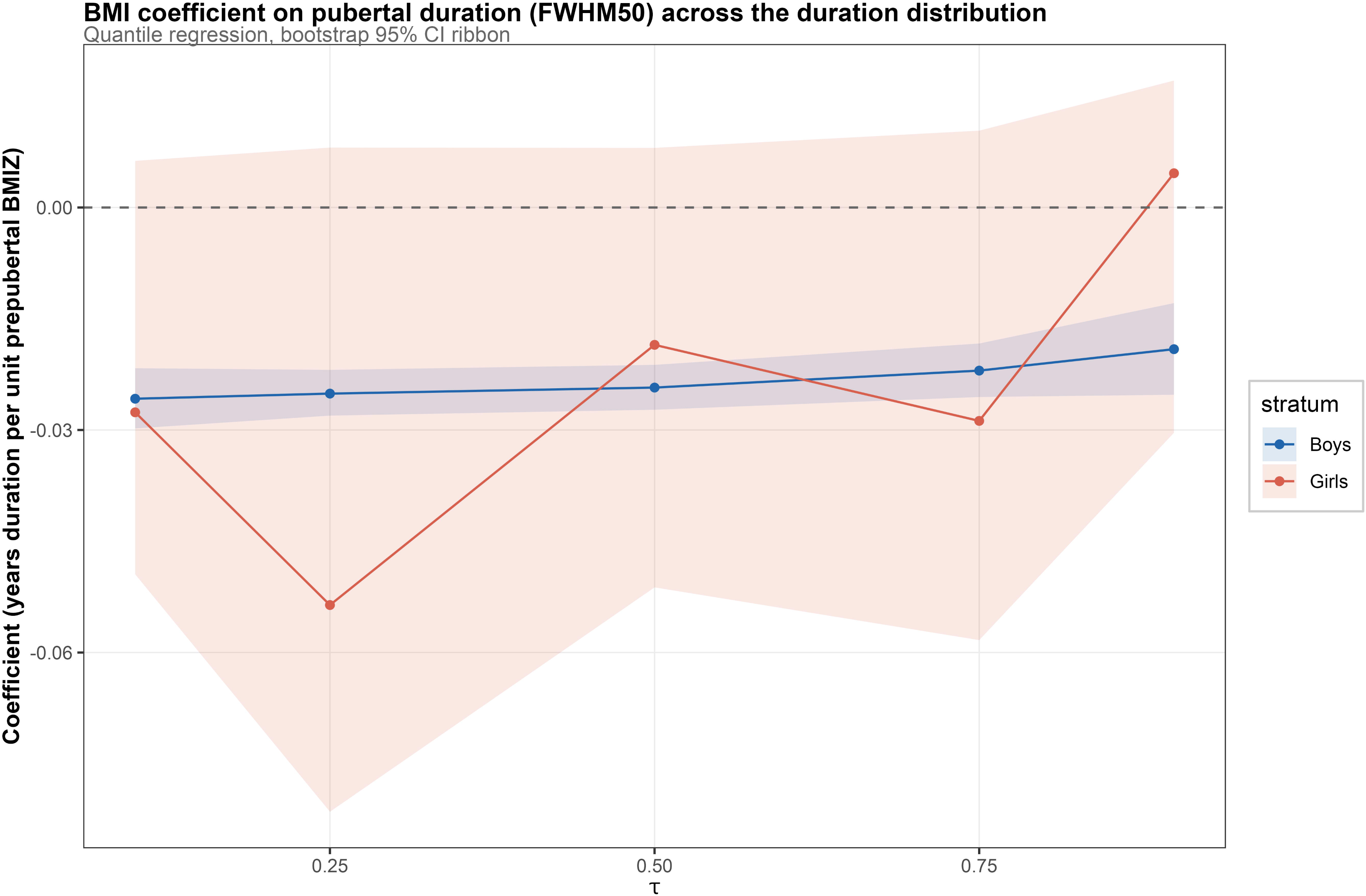
Body mass index coefficient on pubertal duration (full width at half maximum) across the duration distribution. Quantile regression coefficients for prepubertal body mass index z score predicting the full width at half maximum duration measure, estimated separately at five points of the duration distribution (10th, 25th, 50th, 75th, and 90th percentile) for boys (blue) and girls (red). Shaded ribbons show 95% bootstrap confidence intervals. The boys’ coefficient is stable and consistently negative across the distribution. The girls’ coefficient is far less stable and has much wider confidence intervals, consistent with the smaller and less representative identifiable sample described the tables.

Sensitivity analyses supported the main findings. The full width at half maximum association held using a looser 75% of peak velocity threshold in both sexes (**Table S4**), though the stricter 25% threshold could not be estimated for either sex because too few children retained a complete crossing. Widening or narrowing the offset window changed the size of the female full width at half maximum sample considerably, from 0 girls at a 3 year window to 1,282 girls at a 6 year window, with unstable coefficients across this range, while the boys estimate was unaffected by window choice (**Table S5**). Excluding each of the three study cities in turn did not materially change either sex coefficient (**Table S6**). Individual velocity curves grouped by duration tertile are shown in **Figure 1** for descriptive comparison.

## Discussion

In this multicity Vietnamese data, higher prepubertal body mass index was associated with a shorter pubertal growth spurt in unadjusted models for both sexes, using both a threshold based and a threshold free duration measure. This matches the one prior cohort we know of that treated duration as its own outcome, a Chinese study that linked a shorter height spurt, measured from growth takeoff to peak velocity, to a higher risk of overweight and obesity by late adolescence ^13^.

Our finding extends that work by showing the same general pattern in a different population and by validating it with a model free functional measure of curve width, which does not depend on any single SITAR parameter or arbitrary threshold.

The more interesting result came after adjusting for pubertal timing and individual peak height velocity. The association weakened sharply in boys and reversed in girls, so that once timing and intensity were held constant, higher body mass index predicted a slightly longer, not shorter, growth quartile duration in girls. This tells us that body mass index does not act on duration purely through its already documented effects on timing and intensity. Something about a higher body mass index appears to reshape the pubertal velocity curve on its own terms, and it does so differently in girls than in boys. A Danish cohort reached a related conclusion from a different angle, finding that pubertal tempo remained linked to body mass index in young adulthood even after accounting for childhood body mass index, arguing against tempo being simply a downstream consequence of childhood weight ^14^. Our result adds that the direction of this independent link can differ by sex, which has not, to our knowledge, been shown before.

Sex specific patterns around the pubertal growth spurt are not new. A Taiwanese cohort found that puberty itself, rather than body mass index, was the dominant driver of growth velocity in girls entering adolescence, suggesting girls’ growth response to adiposity around puberty may follow a different logic than boys’ ^7^. A Swedish cohort showed that children with higher childhood body mass index can still reach similar adult heights as leaner children by trading more prepubertal growth for less pubertal growth ^5^. Our data suggest a further layer to this trade off, in which the pubertal window itself stretches or shortens in a sex specific way once timing and intensity are accounted for.

A key methodological finding here is a cautionary one. The full width at half maximum measure could be computed for only 18% of girls, and those who could be measured had a noticeably lower body mass index than those who could not. This happened because the fitted female growth model, whose reliable age range is narrower than the male model, could not always capture both the rising and falling half maximum crossings for girls with an early age at peak height velocity, a group already enriched for higher body mass index. This is a form of informative missingness that would bias a naive analysis toward the wrong effect in girls. The growth quartile duration measure, which needs only a child’s own velocity curve rather than a crossing on each side, was available for 95% of girls and is the more trustworthy measure for this sex. Anyone applying width based duration measures from SITAR, Superimposition by Translation and Rotation, models to cohorts with a narrow fitted age range should check for this problem directly, since the model’s own translation and rotation logic does not protect against it^8^.

Whether the duration differences we observed are fixed or modifiable is an open question our design cannot answer, but related work offers a hopeful signal. A prospective cohort of children undergoing obesity treatment found that a hampered pubertal growth spurt was not permanent, and that successful treatment normalized growth velocity rather than causing harm ^28^. If duration behaves similarly, it would support treating pubertal growth pattern changes as a marker of current body mass index rather than a fixed consequence of it.

This study has several limitations. It is observational and cannot establish causation. It draws on Vietnamese schoolchildren from three cities, which may not generalize to rural or lower resource settings. We lacked hormonal, dietary, and socioeconomic data that could help explain the sex difference we found. Strengths include a large sample, sensitivity analyses across cities, thresholds, and window widths, and a functional principal component analysis that confirmed the duration findings using a method independent of the SITAR framework.

These findings carry practical implications for clinicians tracking pubertal growth in children with overweight or obesity. Specifically, how long a growth spurt lasts carries vital information, especially in girls, where higher BMI links to a longer growth window after accounting for timing and intensity. Public health programs in Vietnam and similar countries could leverage growth duration as an early monitoring marker, provided future cohorts track individuals to adult composition. Subsequent research should also explore hormonal factors like leptin or sex steroids to explain sex-specific duration differences across Southeast Asian populations and other regions.

## Data Availability

R codes are available from the corresponding author upon reasonable request. Individual patient-level data cannot be shared due to applicable privacy regulations and the terms of the institutional ethics approval.

## Declaration Contributor’s statement

Nhan Thi Ho did conceptualization, data curation, formal analysis, investigation, methodology, project administration, resources, software, supervision, validation, visualization, writing original draft, and writing review & editing.

## Data Sharing Statement

R codes are available from the corresponding author upon reasonable request. Individual patient-level data cannot be shared due to applicable privacy regulations and the terms of the institutional ethics approval.

## Conflict of intertest

The author states that there is no conflict of interest.

## Use of Artificial Intelligence

The author performed all original research work regarding scientific content, analyses, interpretations and manuscript writing. The author used AI-assisted tools for language editing and grammar checking during manuscript preparation.

