## Supplementary Tables for "Body mass index and the duration of the pubertal growth spurt in Vietnamese urban children"

**Tables and Figure Legends**

*Body mass index and the duration of the pubertal growth spurt in Vietnamese children, a SITAR based functional analysis of velocity curve width*

### **Main Tables**

**Table 1. Cohort characteristics by sex and prepubertal body mass index category**

| **Characteristic** | **Thinness** | **Normal BMI** | **Overweight** | **Obesity** | **Total** | **P value** |
| --- | --- | --- | --- | --- | --- | --- |
| **Boys (n = 8018)** | | | | | | |
| n | 193 | 3613 | 1862 | 2350 | 8018 |  |
| Age at peak height velocity, years | 12.43 (0.50) | 12.30 (0.49) | 12.16 (0.56) | 12.08 (0.58) | 12.20 (0.54) | < 0.001 |
| SITAR timing (b), years | 0.22 (0.50) | 0.09 (0.49) | -0.05 (0.56) | -0.13 (0.58) | -0.01 (0.54) | < 0.001 |
| Individual peak height velocity, cm per year | 9.08 (0.68) | 9.27 (0.65) | 9.43 (0.70) | 9.53 (0.72) | 9.38 (0.69) | < 0.001 |
| Duration, full width at half maximum, years^a^ | 3.43 (0.26) | 3.36 (0.23) | 3.30 (0.24) | 3.26 (0.24) | 3.32 (0.24) | < 0.001 |
| *Missing, n* | *0* | *1* | *1* | *3* | *5* |  |
| Duration, growth quartile, years | 3.66 (0.11) | 3.63 (0.11) | 3.60 (0.13) | 3.59 (0.13) | 3.61 (0.12) | < 0.001 |
| *Missing, n* | *0* | *0* | *0* | *0* | *0* |  |
| City, Hanoi, n (%) | 102 (52.8) | 2398 (66.4) | 1261 (67.7) | 1507 (64.1) | 5268 (65.7) | < 0.001 |
| City, Ho Chi Minh City, n (%) | 76 (39.4) | 885 (24.5) | 454 (24.4) | 606 (25.8) | 2021 (25.2) |  |
| City, Haiphong, n (%) | 15 (7.8) | 330 (9.1) | 147 (7.9) | 237 (10.1) | 729 (9.1) |  |
| **Girls (n = 5491)** | | | | | | |
| n | 179 | 3646 | 1185 | 481 | 5491 |  |
| Age at peak height velocity, years | 9.16 (1.06) | 9.24 (1.00) | 9.27 (0.99) | 9.30 (1.05) | 9.25 (1.01) | 0.420 |
| SITAR timing (b), years | -0.05 (1.06) | 0.03 (1.00) | 0.06 (0.99) | 0.09 (1.05) | 0.04 (1.01) | 0.420 |
| Individual peak height velocity, cm per year | 6.80 (0.64) | 7.00 (0.69) | 7.15 (0.65) | 7.27 (0.70) | 7.05 (0.69) | < 0.001 |
| Duration, full width at half maximum, years^a^ | 6.82 (0.46) | 6.74 (0.45) | 6.70 (0.43) | 6.67 (0.50) | 6.73 (0.45) | 0.320 |
| *Missing, n* | *126* | *2892* | *1038* | *444* | *4500* |  |
| Duration, growth quartile, years | 3.09 (0.18) | 3.04 (0.20) | 2.99 (0.20) | 2.93 (0.27) | 3.02 (0.21) | < 0.001 |
| *Missing, n* | *2* | *145* | *84* | *50* | *281* |  |
| City, Hanoi, n (%) | 105 (58.7) | 2445 (67.1) | 755 (63.7) | 295 (61.3) | 3600 (65.6) | < 0.001 |
| City, Ho Chi Minh City, n (%) | 65 (36.3) | 893 (24.5) | 333 (28.1) | 136 (28.3) | 1427 (26.0) |  |
| City, Haiphong, n (%) | 9 (5.0) | 308 (8.4) | 97 (8.2) | 50 (10.4) | 464 (8.5) |  |

Values are mean (standard deviation) unless otherwise noted. P values compare categories within each sex using the Kruskal Wallis test for continuous variables and the chi square test for city distribution.

BMI, body mass index. SITAR, Superimposition by Translation and Rotation.

a Duration values for girls in this table describe only the subset of girls for whom the full width at half maximum measure could be computed (18 percent of girls overall). This subset is not representative of all girls. See Table S1 and Table S2 for the identifiability pattern and Table 3 for why the growth quartile duration measure is treated as primary for girls.

**Table 2. Correlation of pubertal growth spurt duration with pubertal timing and intensity**

| **Sex** | **Duration with timing, r** | **Duration with intensity, r** | **Timing with intensity, r** | **FWHM with growth quartile duration, r** | **N** |
| --- | --- | --- | --- | --- | --- |
| Boys | 0.85 | -0.99 | -0.85 | 0.95 | 8,013 |
| Girls^a^ | -0.95 | -1.00 | 0.95 | 0.92 | 991 |

Timing is the individual SITAR b parameter. Intensity is each child's own peak height velocity. Duration is the full width at half maximum measure. FWHM, full width at half maximum.

a For girls, these correlations are based on the 991 girls (18 percent) for whom the full width at half maximum measure was identifiable, and should be interpreted with the ceiling limitation described in Table 1 in mind.

**Table 3. Association between prepubertal body mass index and pubertal growth spurt duration**

| **Sex** | **Duration measure** | **N** | **Unadjusted coefficient (95% CI)** | **P value** | **Adjusted coefficient (95% CI)** | **P value** |
| --- | --- | --- | --- | --- | --- | --- |
| Boys | Full width at half maximum | 8,013 | -0.0221 (-0.0257 to -0.0186) | < 0.001 | 0.0004 (0.0000 to 0.0007) | 0.038 |
| Girls^a^ | Full width at half maximum | 991 | -0.0232 (-0.0449 to -0.0014) | 0.037 | -0.0007 (-0.0022 to 0.0007) | 0.330 |
| Boys | Growth quartile duration | 8,018 | -0.0111 (-0.0129 to -0.0094) | < 0.001 | -0.0005 (-0.0010 to 0.0000) | 0.034 |
| Girls | Growth quartile duration | 5,210 | -0.0278 (-0.0329 to -0.0226) | < 0.001 | 0.0052 (0.0017 to 0.0087) | 0.004 |
| **BMI by sex interaction (girls versus boys, reference boys)** | | | | | | |
|  | Full width at half maximum | 9,004 | -0.0018 (-0.0252 to 0.0171) | | 0.824 | |
|  | Growth quartile duration | 13,228 | -0.0166 (-0.0219 to -0.0114) | | 0.007 | |

Coefficients are years of duration per one unit of prepubertal BMI z score, from linear regression with heteroskedasticity consistent standard errors, adjusted for study city. Adjusted models additionally include the individual SITAR timing parameter and individual peak height velocity. The interaction term tests whether the body mass index association with duration differs between girls and boys, with boys as the reference group, using a bootstrap based Wald test.

CI, confidence interval. BMI, body mass index.

a For girls, the full width at half maximum estimate is based on only 991 girls (18 percent of the female sample) because of the identifiability limitation described in Table 1. The growth quartile duration estimate, available for 5,210 girls (95 percent), is the more representative and recommended estimate for girls.

**Table 4. Functional principal component analysis of pubertal velocity curves, robustness check for growth spurt duration**

| **Sex** | **Retained component** | **Correlation with growth quartile duration, r** | **Correlation with full width at half maximum, r** | **Component score regressed on BMI, coefficient** | **P value** | **N** |
| --- | --- | --- | --- | --- | --- | --- |
| Boys | fpc1 | -0.96 | -0.995 | 0.093 | < 0.001 | 8,017 |
| Girls | fpc1 | -0.78 | -0.992 | 0.130 | < 0.001 | 4,629 |

The single retained functional principal component (fpc1) explained more than 99 percent of the variance in the shape of aligned velocity curves within a window of plus or minus 2 years around each child's own age at peak height velocity, for both sexes. Its correlation with the growth quartile duration and full width at half maximum measures shows convergent validity between the model free component score and the two duration measures defined earlier. The final two columns show the component score regressed on prepubertal body mass index z score, with heteroskedasticity consistent standard errors, adjusted for study city.

BMI, body mass index.

### **Supplementary Tables**

**Table S1. Identifiability of the full width at half maximum duration measure, by sex**

| **Sex** | **Full width at half maximum status** | **N** | **Mean prepubertal BMI z score** | **Mean age at peak height velocity, years** | **Obesity, %** |
| --- | --- | --- | --- | --- | --- |
| Girls | Not identifiable | 4,500 | 0.61 | 9.10 | 9.9 |
| Girls | Identifiable | 991 | 0.03 | 9.94 | 3.7 |
| Boys | Not identifiable | 5 | 3.31 | 14.90 | 60.0 |
| Boys | Identifiable | 8,013 | 1.53 | 12.20 | 29.3 |

A child's full width at half maximum duration is identifiable only when the velocity curve crosses 50 percent of that child's own peak velocity on both sides of the peak within the age range supported by the fitted growth model. BMI, body mass index.

**Table S2. Direction of censoring for the full width at half maximum measure and characteristics of censored children**

| **Sex** | **Status** | **N** | **Mean prepubertal BMI z score** | **Mean age at peak height velocity, years** | **Obesity, %** |
| --- | --- | --- | --- | --- | --- |
| Girls | Censored, both sides | 444 | 0.37 | 7.92 | 6.5 |
| Girls | Censored, pre-peak side only | 3,513 | 0.78 | 9.40 | 11.5 |
| Girls | Censored, post-peak side only | 542 | -0.33 | 8.07 | 1.8 |
| Girls | Censored, other or insufficient data | 1 | 2.18 | 19.37 | 0.0 |
| Girls | Identifiable | 991 | 0.03 | 9.94 | 3.7 |
| Boys | Censored, post-peak side only | 5 | 3.31 | 14.90 | 60.0 |
| Boys | Identifiable | 8,013 | 1.53 | 12.20 | 29.3 |

Pre-peak censoring means the curve does not drop to 50 percent of peak velocity before the peak within the available age range. Post-peak censoring means the same on the other side of the peak. Girls censored on the pre-peak side, the largest censored group, have a mean prepubertal BMI z score nearly 0.8 units higher and an obesity prevalence about three times higher than girls with an identifiable duration, indicating that missingness in this measure is related to body mass index and is not random. BMI, body mass index.

**Table S3. Quantile regression of pubertal duration on prepubertal body mass index across the duration distribution**

| **Group** | **Duration measure** | **Quantile** | **Coefficient (95% bootstrap CI)** | **Wald P for heterogeneity across quantiles** |
| --- | --- | --- | --- | --- |
| Boys | Full width at half maximum | 0.10 | -0.026 (-0.030 to -0.022) | 0.313 |
| Boys | Full width at half maximum | 0.25 | -0.025 (-0.028 to -0.022) |  |
| Boys | Full width at half maximum | 0.50 | -0.024 (-0.027 to -0.021) |  |
| Boys | Full width at half maximum | 0.75 | -0.022 (-0.026 to -0.018) |  |
| Boys | Full width at half maximum | 0.90 | -0.019 (-0.025 to -0.013) |  |
| Girls | Full width at half maximum | 0.10 | -0.028 (-0.049 to 0.006) | 0.066 |
| Girls | Full width at half maximum | 0.25 | -0.054 (-0.081 to 0.008) |  |
| Girls | Full width at half maximum | 0.50 | -0.019 (-0.051 to 0.008) |  |
| Girls | Full width at half maximum | 0.75 | -0.029 (-0.058 to 0.010) |  |
| Girls | Full width at half maximum | 0.90 | 0.005 (-0.030 to 0.017) |  |
| Boys | Growth quartile duration | 0.10 | -0.015 (-0.018 to -0.013) | < 0.001 |
| Boys | Growth quartile duration | 0.25 | -0.013 (-0.014 to -0.011) |  |
| Boys | Growth quartile duration | 0.50 | -0.011 (-0.012 to -0.010) |  |
| Boys | Growth quartile duration | 0.75 | -0.009 (-0.011 to -0.008) |  |
| Boys | Growth quartile duration | 0.90 | -0.007 (-0.010 to -0.005) |  |
| Girls | Growth quartile duration | 0.10 | -0.037 (-0.043 to -0.028) | < 0.001 |
| Girls | Growth quartile duration | 0.25 | -0.035 (-0.041 to -0.028) |  |
| Girls | Growth quartile duration | 0.50 | -0.028 (-0.033 to -0.024) |  |
| Girls | Growth quartile duration | 0.75 | -0.020 (-0.024 to -0.017) |  |
| Girls | Growth quartile duration | 0.90 | -0.016 (-0.020 to -0.013) |  |

Coefficients are years of duration per one unit of prepubertal BMI z score at each quantile (tau) of the duration distribution, adjusted for study city, with 95 percent confidence intervals from 300 city stratified bootstrap resamples. The Wald test evaluates whether the coefficient differs across quantiles within each group and duration measure.

BMI, body mass index.

**Table S4. Sensitivity of the body mass index and duration association to the full width at half maximum threshold**

| **Sex** | **Threshold** | **N** | **Coefficient (95% CI)** | **P value** |
| --- | --- | --- | --- | --- |
| Boys | 25% of peak velocity | 0 | Not estimable, too few complete cases | n.a. |
| Girls | 25% of peak velocity | 6 | Not estimable, too few complete cases | n.a. |
| Boys | 75% of peak velocity | 8,016 | -0.0130 (-0.0151 to -0.0109) | < 0.001 |
| Girls | 75% of peak velocity | 4,810 | -0.0322 (-0.0396 to -0.0247) | < 0.001 |

The primary analysis used the 50 percent of peak velocity threshold, reported in Table 3. This table repeats the same model at a stricter (25 percent) and a looser (75 percent) threshold. The 25 percent threshold could not be estimated for either sex because too few children had a curve that dropped this far from peak within the available age range.

n.a., not applicable.

**Table S5. Sensitivity of the body mass index and duration association to the offset window width**

| **Requested window, plus or minus years** | **Sex** | **N** | **Coefficient (SE)** | **P value** |
| --- | --- | --- | --- | --- |
| 3 | Boys | 8,012 | -0.0222 (0.0018) | < 0.001 |
| 3 | Girls | 0 | Not estimable, too few complete cases | n.a. |
| 4 | Boys | 8,013 | -0.0221 (0.0018) | < 0.001 |
| 4 | Girls | 434 | 0.0015 (0.0065) | 0.814 |
| 5 | Boys | 8,013 | -0.0221 (0.0018) | < 0.001 |
| 5 | Girls | 991 | -0.0232 (0.0111) | 0.037 |
| 6 | Boys | 8,013 | -0.0221 (0.0018) | < 0.001 |
| 6 | Girls | 1,282 | -0.0486 (0.0124) | < 0.001 |

Each row repeats the primary full width at half maximum model (Table 3) after rebuilding the velocity panel with a wider or narrower requested window around each child's own age at peak height velocity. The true achievable window remains bounded by each child's sex specific age range regardless of the value requested. SE, standard error. n.a., not applicable.

**Table S6. Leave one city out sensitivity analysis**

| **Sex** | **City excluded** | **Coefficient, full width at half maximum** |
| --- | --- | --- |
| Boys | Hanoi excluded | -0.0176 |
| Girls | Hanoi excluded | -0.0175 |
| Boys | Ho Chi Minh City excluded | -0.0235 |
| Girls | Ho Chi Minh City excluded | -0.0290 |
| Boys | Haiphong excluded | -0.0228 |
| Girls | Haiphong excluded | -0.0222 |

Each row repeats the primary full width at half maximum model (Table 3) after excluding all children from one of the three study cities in turn, to check that no single city drives the overall association.
